# Health facility assessments of cervical cancer prevention, screening, and treatment services in Gulu, Uganda

**DOI:** 10.1101/2022.06.24.22276873

**Authors:** Tana Chongsuwat, Aaliyah O Ibrahim, Ann Evensen, James H Conway, Margaret Zwick, William Oloya

**Affiliations:** Department of Family Medicine and Community Health, University of Wisconsin-Madison, School of Medicine & Public Health, Madison, Wisconsin, USA; Gulu Women’s Economic Development & Globalization (GWED-G), Gulu, Uganda; Department of Pediatrics, University of Wisconsin-Madison, School of Medicine & Public Health, Madison, Wisconsin, USA

## Abstract

**Background:** Cervical cancer is ranked globally in the top 3 cancers for women younger than 45 years, with the average age of death at 59 years of age. The highest burden of disease is in low- and low-to-middle income countries (LLMICs), responsible for 90% of the 311,000 cervical cancer deaths in 2018. This growing health disparity is due to the lack of quality screening and treatment programs, low human papillomavirus (HPV) vaccination rates, and high human immunodeficiency virus (HIV) co-infection rates. To address these gaps in care, we need to develop a clear understanding of the resources and capability of LLMIC’s health care facilities to provide prevention, screening, and treatment for cervical cancer.

**Objectives:** This project aimed to (1) develop a health facility assessment (HFA) to assess available cervical cancer prevention, screening, and treatment resources and (2) implement the HFA to determine the cervical cancer resources available in Gulu, Uganda.

**Methods:** We adapted the World Health Organization’s Harmonized Health Facility Assessment for our own HFA and grading scale, deploying it in October 2021 to analyze 21 health centers in Gulu.

**Results:** Grading of Health Center IIIs (n=16) concluded that 37% had “excellent” or “good” resources available, and 63% of facilities had “poor” or “fair” resources available. Grading of Health Center IVs and above (n=5) concluded that 60% of facilities had “excellent” or “good” resources, and 40% had “fair” resources available.

**Discussion:** The analysis of health facilities in Gulu demonstrated subpar resources available for cervical cancer prevention, screening, and treatment. Focused efforts are needed to expand health centers’ resources and capability to address rising cervical cancer rates and related health disparities in LLMICs. The development process for the study HFA can be applied to global cervical cancer programming to determine gaps in resources and indicate areas to target improved health equity.

## Introduction

Globally, cervical cancer is one of the top 3 cancers in women younger than 45 years, with an average age of death at 59 years of age. The greatest disease burden is in low- and low-to-middle-income countries (LLMICs), contributing to 90% of the 341,000 cervical cancer deaths globally in 2020 [1]. About 12% of new cases occur in African women, yet 85% of deaths occur in Sub-Saharan Africa [2].

Uganda has one of the largest documented cervical cancer disease burdens in Africa, with an incidence rate of 47.5 out of 100,000 women and a mortality rate of 40 out of 100,000 women [3]. In Uganda, the Kampala Cancer Registry collected and calculated incidence rates for different cancers from 1991–2006 and found cervical cancer to be the most common malignancy in women with a 3% annual increase in incidence over 16 years [4]. One half (50.1%) of all female cancers in the Acholi Sub-region, which encompasses Northern Uganda, are cervical, with the most common cases falling between 30 and 49 years and incidence of 57 out of 100,000 women [5].

There are many common challenges for cancer screening and treatment programs in LLMICs, including lack of infrastructure, non-surgical treatment modalities, and chronic shortages of health workers and resources. Additionally, weak referral processes and inadequate health information systems make it difficult to track individual patients or monitor program performance [6,7].

Despite many national health systems prioritizing quality cervical cancer screening programming, these expanded services are significantly underutilized. In Uganda, the health system was decentralized with the intent to improve access and the quality of health services. While this has led to increased utilization of health facilities, it has divided the system into national and district health systems, facing challenges due to pharmaceutical drug shortages, inefficient utilization of resources, and low morale among hospital staff limit implementation and sustainment of cancer programming [8].

In 2010, the Uganda Ministry of Health developed a Strategic Plan for Cervical Cancer Prevention and Control (2010–2014), with priority areas in vaccination against human papilloma virus (HPV), low-cost screening using visual inspection with acetic acid (VIA), and treatment of early dysplasia (cervical intraepithelial neoplasia) using cryotherapy. Expectations in the Strategic Plan on facilities were distinguished based on the type of facility, determined by the target population and the services provided. Facilities are designated as either a health center (levels I to IV, with village health teams designed as level I and serving the smallest target population size) or hospital (general, regional, or national level). In the Strategic Plan, Health Center IIIs (H/C IIIs) and above are expected to provide HPV vaccination and screening by VIA. In addition to vaccination and screening services, Health Center IVs (H/C IVs) and hospitals are expected to provide treatment for early cervical dysplasia [9].

Improving quality services in Northern Uganda for cervical cancer prevention, screening, and treatment is essential. The purpose of this study is to develop and conduct a health facility assessment (HFA) to evaluate the cervical cancer resources of Health Centers III and above in Gulu, Uganda. HFAs are often used to gather large-scale data on a country’s health service availability and quality, generally administered by trained facilitators and may include materials inventory, interviews with patients and staff, and service observation [10]. Results from this assessment in Gulu will inform potential targeted interventions to fill the gaps in cervical cancer prevention, screening, and treatment. Moreover, our HFA development process may inform similar processes for global cervical cancer programming to ultimately counteract the increasing disparities in morbidity and mortality experienced by women in LLMICs.

## Methods

The study was conducted in Gulu, the second-largest city in Uganda, located in the Northern region. The city has 23 health facilities designated as H/C III and above.

### Development of a health facility assessment

In Uganda, facilities are commonly evaluated through the Results-Based Financing program using the Health Center Quarterly Quality Assessment Tool (HCQQAT). Facilities in Uganda are assessed by level, with higher-level facilities expected to provide more services than lower-level facilities. The score a facility receives on this assessment determines the amount of supplementary funding allotted to it and thus incentivizes the provision of services and good healthcare outcomes. The Results-Based Financing approach has been found to improve service coverage by at least 27%, especially for children and pregnant women [11,12].

The World Health Organization (WHO), in their evaluation of HFAs, determined that incongruity between assessment designs has limited cross-assessment comparisons of health facilities. Therefore, the WHO created the Harmonized Health Facility Assessment (HHFA), a more comprehensive questionnaire designed to be administered either as a whole or as individual modules [13]. We developed a modified HFA using assessment questions from the WHO HHFA cervical cancer questionnaire that was then adapted to the style of the Ugandan HCQQAT to make it contextually relevant for the Ugandan field officers. The modified HFA included questions in 3 sections: (1) services available, such as the ability to perform certain services related to screening, treatment, and vaccination; (2) facility support for service performance, such as providers trained to perform VIA, HPV diagnostic testing, colposcopy, and excisional or ablative treatments; and (3) available and functioning equipment for cervical cancer services.

### Data collection and analysis

Local approval was obtained from the District Health Officer prior to delivery of the HFA. Institutional review was not required, as this project was considered a program evaluation and not a human-subjects research project. Gulu Women’s Economic Development & Globalization, a women-led, grassroots organization focused on gender equality, selected field officers with experience gathering assessments from targeted facilities. They received training on cervical cancer and how to administer this HFA, then collected data in October 2021 from all 23 health facilities listed as Health Center III and above. Field officers prioritized gathering data from the facility chair or a supervisor of the maternity ward or outpatient department, depending on availability.

The HFA used in this study scored each section with points given for available resources. Because there are different levels of expectations based on a facility level, it was difficult to determine if a health facility had adequate resources based on this score alone. Thus, a grading system (Table 1) was developed to determine if facilities met the expectations outlined by the Uganda Ministry of Health’s Strategic Plan. Facilities that reported the ability to provide VIA screening but without acetic acid in stock were not counted as having the resource available. Similarly, for the equipment portion of the assessment, field officers requested to check functionality of equipment such as the colposcope, loop electrosurgical excisional procedure (LEEP) machine, cryotherapy, or other materials. If the equipment was not functioning, it was determined the facility did not have the service available. Facilities were determined to have adequate support for pelvic examination if specific equipment was available, including speculums, gloves, and a dedicated gynecological exam table with functioning stirrups. For example, a Health Center III that was able to provide both HPV vaccination and screening with VIA or another method but did not have a dedicated gynecological exam table with stirrups received a “good” grade, as more equipment support was needed to comfortably perform a pelvic examination.

**Table 1.** Grading of Health Facilities based on Strategic Plan Expectations.

| **Grading** | **Health Center III** | **Health Center IV and Above** |
| --- | --- | --- |
| **Excellent** | Able to provide both services (HPV vaccine and screening) | Provides all 3 with adequate support |
| **Good** | Able to provide both services (HPV vaccine and screening) but more support needed | Provides all 3 services but more support needed |
| **Fair** | Only able to provide 1 service | Able to provide 2 of 3 services |
| **Poor** | Unable to provide either HPV vaccines or screening | Not able to provide any services or only 1 |

## Results

A summary of the 21 facilities that responded to the HFA (16 H/C IIIs, 1 H/C IV, and 4 hospitals) is reported as simple statistics. Two facilities declined to provide data. HPV vaccination was available at 65% (n=13) facilities. Screening (VIA, pap smear, and/or HPV testing) was possible at 71% (n=15) facilities. Early treatment for cervical dysplasia was available at 6 locations, including 4 of the 5 facilities where treatment services were expected (H/C IVs and above) and 2 H/C IIIs. Only 1 facility was able to provide excisional treatment through LEEP and reported functioning equipment except for a smoke evacuator. Since the procedure could still safely be performed without this equipment, excisional treatment was included in the resource list for this facility. A summary of resources available by health facility level is included in Table 2.

**Table 2.** List of Resources Available by Facility Level.

|  | Health Center<br>III<br>(n = 16) |  | Health Center IV<br>and Hospitals<br>(n = 5) |  | **Total**<br>**(n = 21)** |  |
| --- | --- | --- | --- | --- | --- | --- |
|  | n | % | n | % | n | % |
| **Prevention** |  |  |  |  |  |  |
| Vaccination for HPV | 9 | 56 | 4 | 80 | 13 | 62 |
| **Screening** |  |  |  |  |  |  |
| Pap Smear (Cytology) | 3 | 19 | 2 | 40 | 5 | 24 |
| Visual Inspection with Acetic Acid (VIA) | 10 | 63 | 5 | 100 | 15 | 71 |
| HPV PCR testing | 0 | 0 | 2 | 40 | 2 | 10 |
| **Treatment** |  |  |  |  |  |  |
| Ablation (Cryotherapy) | 2 | 13 | 4 | 80 | 6 | 29 |
| Excisional (LEEP <sup>a</sup>) | 0 | 0 | 1 | 20 | 1 | 5 |
| **Other Support Needs** |  |  |  |  |  |  |
| Gynecological Bed | 8 | 50 | 4 | 80 | 12 | 57 |
| Stirrups | 4 | 25 | 3 | 60 | 7 | 33 |
| Biopsy | 0 | 0 | 3 | 60 | 3 | 14 |
| Colposcopy | 0 | 0 | 2 | 40 | 2 | 10 |

<sup>a</sup> Loop electrosurgical excision procedure

Grading of H/C III facilities (Figure 1) concluded that 37% had “excellent” or “good” resources available, and 63% of facilities had “fair” or “poor” resources available. Grading of facilities H/C IV and above (Figure 2) concluded that 60% had “excellent” or “good” resources, and 40% of facilities had “fair” resources available.

**Figure 1.**
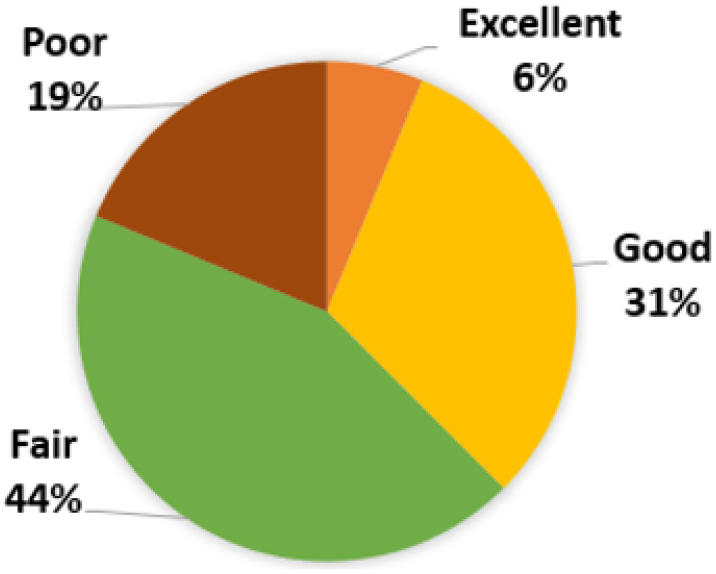
Grading of Health Center III Cervical Cancer Service.

**Figure 2.**
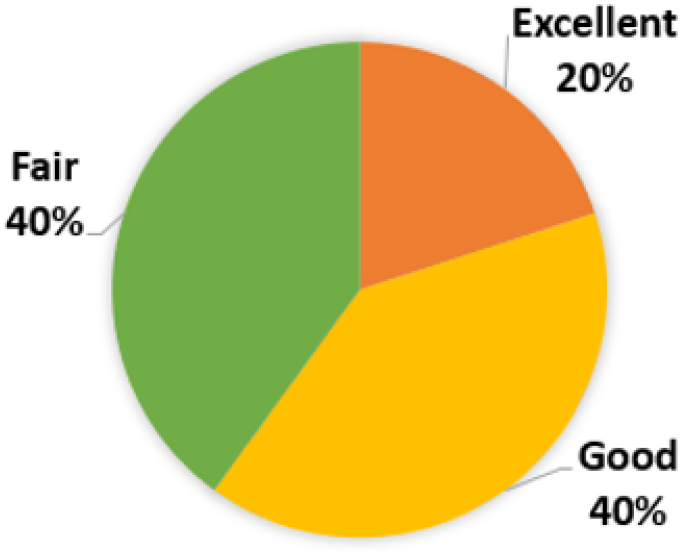
Grading of Health Center IV and Above Cervical Cancer Service.

## Discussion

Efforts to build capacity for cervical cancer control in LLMICs are challenged by financing, health care worker shortages, institutional bias, and limitations due to the ongoing COVID-19 pandemic. Current roadmaps and evidence-based interventions call for expanded capacity-building efforts, particularly HPV vaccination and same-day screen-and-treat approaches [14,15].

HPV vaccination in Uganda first became available in 2006, with nationwide rollout of an HPV vaccination program starting in 2015 and including a goal of achieving 80% coverage for eligible girls. As of 2020, an estimated 74% of eligible girls have received at least 1 dose, and 30% of girls have completed the series [1]. Various individual and community factors impact vaccination uptake, with correlation between series completion and older age, higher attendance at a school-based program, higher socioeconomic status (SES), and living in an urban location [16]. While this study and HFA evaluated the access to HPV vaccines through health facilities, the national vaccination program focused primarily on girls age 10 to 14 in schools and community outreach program, therefore the HFA may not accurately capture the vaccine accessibility in the country.

Cervical cancer screening methods include VIA followed by cytology and, most recently, HPV polymerase chain reaction (PCR) testing. Compared to HPV PCR testing, VIA can be less sensitive but more specific for detecting cervical intraepithelial neoplasia 2+. VIA sensitivity and specificity is approximately 80% (95% CI 0.79-0.82) and 92% (95% CI 0.91-0.92), respectively, compared to HPV PCR Hybrid Capture II (Qiagen, Germany) sensitivity and specificity of 84.0% (95% CI 0.742–0.906) and 88.3% (95% CI 0.818–0.927), respectively [17,18]. Cytology requires pathologists and/or pathology technicians to examine samples, costly services that are often physically remote from the health center performing the screening. HPV PCR testing is expensive and not widely available. Both cytology and HPV PCR testing require follow-up at a second visit for treatment. Results in this study demonstrate a higher availability of VIA as a screening method in facilities evaluated. Focus for cervical cancer programs in similar settings should support VIA as a low cost and accurate screening method. Training for VIA can be done in 2 weeks for nurses and non-physician health care provider, requires very little resources, and promotes screen-and-treat approaches in LLMICs.

Vaccination and early screening through VIA have significantly reduced morbidity and mortality, providing up to 97% relative reduction in cervical cancer rates for girls aged 12–13 years old compared to unvaccinated individuals [19]. Additionally, screening at the age of 35 years using a one- or two-visit screening strategy involving VIA has been found to reduce the risk of cervical cancer by approximately 25% to 36% [20].

To support these successful prevention strategies, this project was developed to evaluate programs in Gulu, Uganda and provide a systems map for health care workers, public health organizations, and community members. The data catalogs the resources available at individual facilities and highlights gaps in resources at facilities that are expected to provide services related to cervical cancer (H/C IIIs and above).

Limitations of this project include some confusion and inconsistency in responses to the HFA used, which suggests a need to re-evaluate certain questions and perform further validation studies for the tool. For example, one item included “Read HPV test,” from the WHO standardized HHFA which was commonly misinterpreted by respondents because it was unclear if this meant ability to run HPV PCR testing on laboratory diagnostic equipment, interpret HPV results, or detect HPV dysplastic changes on the cervix (through VIA, DC, or colposcopy). Based on the Uganda Ministry of Health’s Strategic Plan, expectations of facilities’ ability to provide certain services differed by facility type, but this study used the same HFA for all facilities. Due to these differing expectations, grading proved to be more nuanced than the calculated score. For example, a hospital with an assessment score of 26 points may score “poor” overall due to mismatch between available and expected treatment services, while a health center with an “excellent” score may receive an assessment score of 19 points for providing all basic resources expected of their facility.

Finally, this HFA evaluated resources available but does not evaluate if health care workers actually provided screening or treatment services to patients presenting to health centers needing them. A recent survey of health care workers at H/C IIIs and H/C IVs in Northern Uganda found that only 18% reported being able to conduct screening, and 57% reported relying on outreach from outside organizations for screening, indicating a possible disconnect between resource availability and utilization when indicated [21].

## Conclusion

Our analysis of health facilities in Gulu, Uganda was performed through a modified health facility assessment incorporating assessment questions from the WHO Harmonized Health Facility Assessment tool and utilizing local expertise to perform the survey. Results from 21 of the 23 facilities demonstrated subpar resources available in the facilities expected to provide cervical cancer prevention, screening, and treatment services in their communities. Focused efforts on expanding health centers’ resources and capabilities to sustainably provide services will be essential in reducing cervical cancer rates and related health disparities in LLMICs.

## Data Availability

Data included in the supplementary files

## Acknowledgements

Authors would like to acknowledge Rachel Lundwall of University of Wisconsin-Madison for providing writing assistance and Francis Okongo of Gulu, Uganda for feedback on the data collection tools.

